# Accelerometry-assessed sleep duration and timing in late childhood and adolescence in Scottish schoolchildren: a feasibility study

**DOI:** 10.1101/2020.04.06.20055434

**Authors:** Laura M. Lyall, Natasha Sangha, Cathy Wyse, Elaine Hindle, Dawn Haughton, Kate Campbell, Judith Brown, Laurence Moore, Sharon A. Simpson, Joanna C. Inchley, Daniel J. Smith

**Affiliations:** Mental Health and Wellbeing, Institute of Health and Wellbeing, University of Glasgow, UK; Institute of Biodiversity, Animal Health and Comparative Medicine, University of Glasgow, UK; School of Biomolecular and Biomedical Science, University College Dublin, Ireland; Medical Research Council/Chief Scientist Office Social and Public Health Sciences Unit, University of Glasgow, UK

**Keywords:** sleep, circadian, adolescence, children, depression, mental health

## Abstract

Children and adolescents commonly suffer from sleep and circadian rhythm disturbances, which may contribute to poorer mental health and wellbeing during this critical developmental phase. Many studies however rely on self-reported sleep measures. This study assessed whether accelerometry data collection was feasible within the school setting as a method for investigating the extent of sleep and circadian disruption, and associations with subjective wellbeing, in Scotland. Fourteen days of wrist-worn accelerometry data were collected from 69 pupils, aged 10-14 years. Objective measures of sleep timing, sleep duration and circadian rest-activity patterns were derived. Questionnaires assessed subjective sleep timing, depressive symptoms, and experiences of wearing the accelerometer. Pupils slept on average less than 8 hours per night, failing to meet standard age-specific recommendations. Sleep timing was later and duration longer on weekends compared to weekdays (B = 0.87, 95% confidence interval (CI) 0.70, 1.04; B = 0.49, 95% CI 0.29, 0.69), indicating social jetlag. Lower daytime activity was correlated with higher depressive symptoms (*r* = -0.84, *p* = 0.008). Compared to primary school pupils, secondary pupils had shorter sleep window duration and lower circadian relative amplitude. Over half of participants reported some discomfort/inconvenience wearing the accelerometer. These data highlight that inadequate sleep is prevalent in this sample of schoolchildren. Future, larger scale investigations will examine in more detail the associations between sleep, circadian function and physical activity with mental health and wellbeing.

## Introduction

Sufficient high-quality sleep and robust 24 hour circadian rhythms (recurring patterns in behavioural and physiological processes including rest-activity patterns) are critical for mental and physical wellbeing (Foster et al., 2013; Roenneberg & Merrow, 2016). Chronic sleep and circadian disturbances have, for example, been repeatedly associated with greater risk of mood disturbances and with cardiometabolic ill health (Cappuccio, Cooper, Delia, Strazzullo, & Miller, 2011; McClung, 2013). Late childhood and adolescence represents a critical period when sleep and circadian rhythms are often disturbed (Touitou, Touitou, & Reinberg, 2016), and when onset of mental illness frequently occurs (Lee et al., 2014); the two being directly linked in some contexts (Lovato & Gradisar, 2014).

From around 10 years of age until late adolescence, delays in the timing of the circadian clock lead to increased preference for evening versus morning activity (an evening chronotype), alongside later preferred timing of sleep (Randler, Faßl, & Kalb, 2017; Roenneberg et al., 2004). Increased propensity towards evening chronotype is a key contributing factor in sleep disturbances in late childhood and adolescence, and is associated with later sleep onset, shorter sleep duration, poor quality sleep and daytime sleepiness (Giannotti, Cortesi, Sebastiani, & Ottaviano, 2002). Computer and social media use before bed further affects circadian rhythms and delays sleep onset in many schoolchildren (Harbard, Allen, Trinder, & Bei, 2016). Exposure to artificial light at night, through lighting both at home and from streetlights, can exacerbate the influence of later chronotype by further delaying the circadian clock (Touitou et al., 2016).

Such factors contribute to widespread prevalence of insufficient and poor-quality sleep in young people. The American Academy of Sleep Medicine recommends that for optimal mental and physical health, children aged 6-12 years should regularly sleep 9-12 hours per night, and teenagers 8-10 hours (Paruthi et al., 2016). These targets are often not met. Of almost 25,000 Canadian children and teenagers surveyed in 2013/2014 (Chaput & Janssen, 2016), nearly a third of 10-13 year olds and over a quarter of 13-17 year olds did not meet these guidelines. A later US study of over 50,000 school students found that 58% of 9-13 year-olds and 73% of 13-18 year olds reported insufficient sleep duration (Wheaton, Jones, Cooper, & Croft, 2018). Smaller studies, primarily using self-report measures, suggest that short sleep duration similarly affects European populations (Ghekiere et al., 2019). Difficulty falling asleep and poor sleep efficiency are prevalent in schoolchildren in many countries (Spruyt, O’Brien, Cluydts, Verleye, & Ferri, 2005). Desynchrony between an individual’s chronotype and the sleep-wake schedule enforced by school timing (*social jetlag*) can lead to sleep problems being particularly common on school nights compared to weekends (Wittmann, Dinich, Merrow, & Roenneberg, 2006).

Disturbed sleep and circadian misalignment in young people are associated with worse school and cognitive performance (Dewald, Meijer, Oort, Kerkhof, & Bögels, 2010), as well as behavioural and psychiatric problems: mood disturbances and depressive symptoms seem to be particularly common in children and adolescents with sleep/circadian disturbances (Lovato & Gradisar, 2014). Given that most psychiatric disorders begin in childhood and adolescence, it is of great importance to clarify the role of sleep and circadian health in mental health. For example, recent meta-analytic evidence suggests that sleep disturbances in adolescents often precede the onset of depression (Lovato & Gradisar, 2014).

While the prevalence of sleep and circadian disturbances and their impact on mental wellbeing is increasingly well established in adolescents and in early childhood, surprisingly few studies have assessed the extent of this issue in late childhood (i.e. 9-12 years), or within the British population. Further, most assessments of sleep in child and adolescent populations have relied on self-report measures from young people or their parents, despite growing evidence of a lack of concordance between self-report and objective (accelerometry) measures of sleep characteristics (Aili, Åström-Paulsson, Stoetzer, Svartengren, & Hillert, 2017). Wrist-worn accelerometry permits non-invasive objective measurement of sleep, circadian and activity characteristics, while participants continue their normal daily activities.

In Scotland, the Schools Health and Wellbeing Improvement Research Network (SHINE; https://shine.sphsu.gla.ac.uk/), provides an established infrastructure for schools-based research and takes a data-driven, systems-level approach to mental health improvement in schools. The current membership includes 169 schools. A key aim of the network is to develop engagement of schools with health and wellbeing research: the importance of sleep in children and adolescents has been identified as a key research priority for schools, with many expressing willingness to participate in such research. Working with four collaborating schools in Central Scotland, we assessed the feasibility and acceptability of accelerometry-based measures to examine the prevalence of sleep/circadian disturbance, and its associations with mental health and wellbeing. As well as investigating the extent of sleep and circadian disturbances, we aimed to assess response rates and data completeness, and any barriers to data collection. The findings will inform the design of a larger-scale study investigating the role of sleep in mental wellbeing in young people.

## Methods

### Participants and procedure

This study was approved by the University of Glasgow College of Medical, Veterinary & Life Sciences Ethics Committee, and written informed consent was obtained from both participants and a parent/legal guardian. One hundred pupils from four SHINE Network schools in Central Scotland were invited to participate: parental and pupil consent was obtained from 74 pupils. Participating schools were offered feedback of pseudonymised, school-level results.

To allow examination of associations of socioeconomic deprivation with sleep characteristics, one primary and one secondary school were in areas of high socioeconomic deprivation, defined as 40-45% of pupils living within Scotland’s most deprived datazones, i.e. the highest Scottish Index of Multiple Deprivation (SIMD) quintile. The remaining primary and secondary schools were in the least deprived areas, with 0-15% of pupils living in the highest SIMD quintile areas.

Data collection took place in October and November 2019. Participants were issued with a wrist-worn triaxial accelerometer (Axivity AX3) by a fieldworker who visited each school. Participants were asked to wear the accelerometer on their dominant wrist continuously during all normal activities for 14 days, removing it only if it caused discomfort, or when submerged in water (i.e. swimming/bathing). Data recording commenced around 2 hours after the school visit (start time from 10 am – 2.15 pm). After 14 days, the accelerometers were collected by the same fieldworker, and participants were asked to complete a demographic information form, questionnaires on their sleep and wellbeing over the preceding two weeks, and to give feedback on their experience of wearing the accelerometer (see *Questionnaires*). Repeat visits to collect accelerometers from pupils absent on the day of collection were necessary for two schools.

Due to omissions in demographics recording, sex was not recorded, and only five participants reported their age. Where age was not reported, this was estimated based on the typical age of pupils in each year group at the start of the school year (as data were collected in October/November):11 for Primary 7 (n = 30), 12 for S1 (n = 24), and 14 for S3 (n = 7). Sex was determined based on first names provided by participants. Where names were unisex and/or uncommon (n = 9), National Records of Scotland archives were examined for the probable year of birth of the relevant year group: numbers of male and female babies given the name were compared, and where a clear majority (>68%) were of one sex, this was assigned. Sex was coded as missing for one participant whose first name did not appear in NRS records, and one further participant whose first name did not show a clear sex-specific majority for their year of birth.

### Accelerometry pre-processing

Accelerometers were configured to record raw acceleration data at a frequency of 50Hz with a range of ± 8g using Open Movement GUI (OMGUI, V1.0.0.42). Binary format accelerometry data (.cwa) were processed in R (v.3.6.1.) using the GGIR package (https://cran.r-project.org/web/packages/GGIR/index.html). Autocalibration was applied: no calibration errors were detected. Metrics of the average magnitude of dynamic acceleration (Euclidean Norm Minus One; ENMO) and arm angle were calculated, and periods of non-wear time were detected based on 15-minute windows (van Hees et al., 2013)

For one primary and one secondary school (n=24 with valid data), the end of daylight savings time (DST) occurred within the first 3-4 days of data collection. Sleep window duration, onset and wake times (see below) were adjusted for this clock change and so were included for all available nights. Measures of sleep duration within the sleep window and sleep efficiency (see *‘Sleep* Measures’) were however inflated due to the DST change, and were therefore coded as missing for the relevant night.

### Activity/circadian rest-activity measures

Using the ENMO metric, average activity during each participant’s least active 5 hours (L5; averaged across each available 24-hour period), and during their most active 10 hours (M10; averaged across 24-hour periods) were calculated. M10 and L5 are commonly used to represent day- and night-time activity, respectively (Witting, Kwa, Eikelenboom, Mirmiran, & Swaab, 1990), and were calculated across all available days, and separately for weekday (Sunday – Thursday) and weekend (Friday – Saturday) nights.

Using the average L5 and M10 values, relative amplitude (RA) was calculated as:

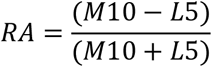

RA reflects the relative difference in activity between the most and least active periods of the day and is a common nonparametric measure of rest-activity rhythm amplitude, ranging from 0-1 (Witting et al., 1990). Lower values can reflect disturbed sleep, sedentary activity during waking hours, or both: higher values indicate ‘healthier’ rest-activity rhythms.

### Sleep measures

The sleep period time (SPT) window (the time window from first sleep onset until last waking of the night, i.e. time in bed) was calculated without sleep diary (van Hees et al., 2018), based on z-angle variance. From this, the average time of sleep onset and waking (time of the start and end of the SPT window, respectively) were calculated. The total duration of sustained inactivity bouts (no change in z-angle of > 5° for at least 5 minutes) within the SPT-window was summed to calculate sleep duration per night, and averaged across available nights (van Hees et al., 2018). Sleep efficiency was defined as sleep duration within SPT divided by the total SPT-window duration, i.e. the proportion of the SPT-window spent asleep. For 24 participants, the night of the end of DST was excluded from calculation of average sleep efficiency and sleep duration values.

### Social jetlag

Based on SPT-window onset and wake times (see above), measures of the difference in sleep timing between weeknights and weekend nights (*social jetlag*) was derived from:

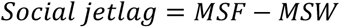

where MSF = the midpoint of sleep on free (weekend) nights (Friday, Saturday), and MSW = the midpoint of sleep on weeknights (Sunday-Thursday). Positive values indicate later sleep timing on weekend nights.

### Questionnaires

At the end of the accelerometry data collection period, participants were asked to complete pencil and paper assessments of their sleep and mental wellbeing during the preceding two weeks, and to evaluate their experience of wearing the accelerometer.

#### Short Adolescent Sleep Wake Scale (ASWS)

The short ASWS (Essner, Noel, Myrvik, & Palermo, 2015) is a 10-item questionnaire assessing subjective sleep quality along three dimensions: going to bed (e.g., ‘*When it’s time to go to bed, I want to stay up and do other things*.’), falling asleep and reinitiating sleep after waking (e.g., ‘*When it’s time to go to sleep, I have trouble settling down*.’), and returning to wakefulness following sleep (e.g., ‘*In the morning, I wake up feeling rested and alert*.’), in addition to a total sleep quality score. Items were scored along a 6-point scale from ‘*Never*’ to ‘*Always’*, with some items reverse coded so that higher scores represent ‘better’ subjective sleep quality.

#### Health Behaviour in School-Aged Children (HBSC): Sleep Questions

The HBSC Sleep Questions asked participants to describe their usual time of going to bed and waking, separately for school nights and weekend/holiday nights. Response options were check boxes at half hourly intervals from ‘*No later than 21*.*00’* until ‘*02*.*00 or later*’ or ‘*04*.*00 or later’* (school days and weekends, respectively) for going to bed; and for waking, from ‘*No later than 05*.*00*’ until ‘*08*.*00 or later*’ for school mornings, and from ‘*No later than 07*.*00*’ until ‘*14*.*00 or later*’ for weekend mornings. Estimates of subjective sleep onset and wake times, and subjective social jetlag (as described above) were calculated.

#### Mood and Feelings Questionnaire (MFQ): Short Version

The MFQ (short version) (Costello & Angold, 1988) assessed frequency of depressive symptoms over the previous two weeks, including items such as ‘*I felt miserable or unhappy*’. Items were rated ‘*Not true*’, ‘*Sometimes’* or ‘*True*’. Scores range from 0 to 26, with higher scores indicating more depressive symptoms.

#### Feedback Assessment

Participants were asked to describe whether they had any problems wearing the accelerometer or completing the questionnaires, and to provide further free text details if responding ‘*yes*’.

## Results

### Sample characteristics and feasibility measures

We aimed to recruit up to 25 pupils per school, i.e. 100 pupils in total (see Table 1). Seventy-four pupils gave consent to participate, with numbers recruited per school ranging from 56% to 100% of the targeted 25. Sixty-nine pupils provided both accelerometry and questionnaire data: 34 from primary schools (Primary 7, aged around 11 years); and 35 from secondary schools (S1/S3, aged 12 – 14 years; Table 1). One further pupil wore the accelerometer but did not complete the questionnaires, and so was excluded. Analyses were restricted to the 61 participants with valid accelerometer data available for at least 1 weekend day, 3 weekdays, and 4 nights: for three schools, valid data was provided by at least 85% of recruited pupils, but for the remaining (secondary) school, less than half of recruited pupils provided valid data.

**Table 1.**
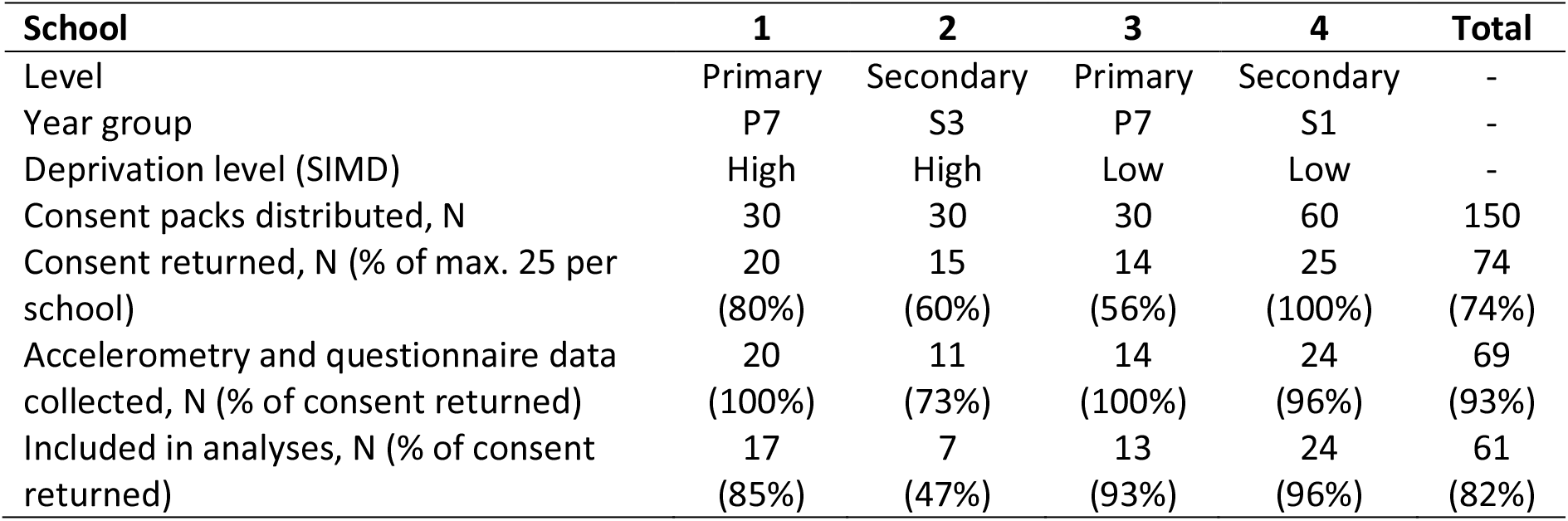
Overview of numbers recruited and included in analyses from each participating school.

| **School** | **1** | **2** | **3** | **4** | **Total** |
| --- | --- | --- | --- | --- | --- |
| Level | Primary | Secondary | Primary | Secondary | - |
| Year group | P7 | S3 | P7 | S1 | - |
| Deprivation level (SIMD) | High | High | Low | Low | - |
| Consent packs distributed, N | 30 | 30 | 30 | 60 | 150 |
| Consent returned, N (% of max. 25 per school) | 20<br>(80%) | 15<br>(60%) | 14<br>(56%) | 25<br>(100%) | 74<br>(74%) |
| Accelerometry and questionnaire data collected, N (% of consent returned) | 20<br>(100%) | 11<br>(73%) | 14<br>(100%) | 24<br>(96%) | 69<br>(93%) |
| Included in analyses, N (% of consent returned) | 17<br>(85%) | 7<br>(47%) | 13<br>(93%) | 24<br>(96%) | 61<br>(82%) |

Table 2 provides a summary and comparison of sample characteristics, accelerometry and questionnaire data for primary (n = 30) and secondary (n = 31) pupils. Included participants provided accelerometry data for the majority of the data collection period (maximum nine weekdays, four weekend days, 14 nights), indicating good compliance with study instructions (Table 2). A greater proportion of primary (0.57) than secondary (0.23) pupils were from more deprived schools. Generalisability of the findings reported below is likely limited due to the small sample size, and limited power may have led to Type 2 error.

**Table 2.** Comparison of sample characteristics between primary and secondary school pupils.

| Measure (M (SD)) | Primary<br>(n=30) | Secondary<br>(n=31) | t/$\chi^2$ | p |
| --- | --- | --- | --- | --- |
| Datasets excluded, N (% of sample with accelerometry) | 4 (11.76) | 4 (11.43) | 0.002 | 0.965 |
| Age*, M (SD) | 10.97 (0.18) | 12.39 (0.92) | 8.30 | **<0.001** |
| Female*, N (%) | 17 (56.67) | 22 (70.97) | 1.43 | 0.490 |
| missing, N (%) | 1 (3.33) | 1 (3.23) |  |  |
| SIMD higher deprivation school, N (%) | 17 (56.67) | 7 (22.58) | 7.42 | **0.006** |
| Number valid weekdays | 8.27 (1.48) | 8.45 (1.34) | 0.51 | 0.611 |
| Number of valid weekend days | 3.60 (0.81) | 3.61 (0.88) | 0.06 | 0.953 |
| Number of nights | 12.73 (2.23) | 13.10 (1.85) | 0.69 | 0.490 |
| Wore accelerometer on dominant wrist, N (%) | 16 (53.33) | 18 (58.06) | 1.10 | 0.577 |
| missing, N (%) | 1 (3.33) | 0 (0) |  |  |
| **Accelerometry-derived activity measures** |  |  |  |  |
| Activity (ENMO) during L5 | 3.03 (0.84) | 2.91 (0.13) | 0.59 | 0.556 |
| Activity (ENMO) during M10 | 76.00 (21.61) | 54.84 (12.37) | 4.71 | **<0.001** |
| **Accelerometry-derived sleep measures** |  |  |  |  |
| SPT-window duration (hours) | 8.81 (0.67) | 8.40 (0.71) | 2.36 | **0.021** |
| Sleep duration (hours) | 7.67 (0.73) | 7.32 (0.64) | 1.96 | 0.054 |
| Sleep efficiency | 0.87 (0.03) | 0.87 (0.04) | 0.19 | 0.854 |
| Sleep onset – weekdays | 22:23 (44.4) | 22:50 (53.8) | 2.20 | **0.032** |
| Sleep onset – weekends | 23:23 (67.2) | 23:36 (75.6) | 0.66 | 0.514 |
| Wake time – weekdays | 07:05 (39.4) | 07:00 (38.6) | 0.51 | 0.610 |
| Wake time - weekends | 08:17 (77.7) | 8:32 (68.3) | 0.80 | 0.425 |
| Meets recommended sleep duration, N (%) | 0 (0) | 1 (3.23) | 0.98 | 0.321 |
| SPT-window meets recommended sleep duration, N (%) | 11 (36.67) | 10 (32.36) | 0.13 | 0.717 |
| **Accelerometry-derived circadian measures** |  |  |  |  |
| Relative amplitude | 0.92 (0.03) | 0.90 (0.03) | 3.17 | **0.002** |
| Social jetlag (hours) | 1.10 (0.91) | 1.14 (0.80) | 0.17 | 0.866 |
| **Questionnaires** |  |  |  |  |
| Total Sleep Score | 35.47 (10.06) | 37.45 (8.22) | 0.85 | 0.402 |
| Falling Asleep & Reinitiating Sleep | 19.20 (6.74) | 21.35 (5.82) | 1.34 | 0.186 |
| Returning to Wakefulness | 5.57 (2.56) | 5.35 (2.27) | 0.34 | 0.733 |
| Going to Bed | 10.70 (3.60) | 10.74 (2.73) | 0.05 | 0.959 |
| Mood and Feelings score | 3.80 (4.45) | 5.35 (3.88) | 1.46 | 0.151 |
| **Subjective sleep/circadian measures** |  |  |  |  |
| Subjective sleep onset – weekdays | 22.06 (8.34) | 22.09 (10.0) | 0.21 | 0.836 |
| Subjective wake time - weekdays | 07:05 (37.8) | 07:09 (31.2) | 0.42 | 0.677 |
| Subjective sleep onset – weekends | 23:41 (98.4) | 23:07 (76.8) | 1.52 | 0.135 |
| Subjective wake time - weekends | 09:27 (95.4) | 09:06 (76.1) | 0.96 | 0.341 |
| Subjective sleep duration – weekdays | 8.98 (1.12) | 9.00 (1.22) | 0.06 | 0.956 |
| Subjective sleep duration - weekends | 9.77 (1.96) | 9.98 (1.57) | 0.48 | 0.633 |
| Subjective social jetlag | 1.98 (1.16) | 1.46 (0.88) | 1.96 | 0.055 |
| **Feedback questions** |  |  |  |  |
| Problems with questionnaires? N (%) | 1 (3.33) | 0 (0) | 1.05 | 0.591 |
| Problems with accelerometer? N (%) | 15 (50.00) | 17 (54.84) | 1.11 | 0.574 |

\* Age and sex are estimated, see *Participants and Procedure*. Values reported in 'Primary' and 'Secondary' columns are mean (standard deviation) unless otherwise specified. For sleep onset and wake times, values are time in 24hr clock (SD in minutes). ENMO = Euclidian Norm Minus One; SPT = Sleep Period Time.

Just over half of included pupils wore the accelerometer on their dominant wrist, as instructed (Table 2). To assess for any impact on results, independent t-tests or chi-square tests of association (for continuous and categorical measures, respectively; full results not presented) compared each accelerometry measure between participants wearing the accelerometer on their dominant and non-dominant wrist, collapsing across primary and secondary pupils. Participants using their dominant wrist had on average fewer nights of valid data (*M* = 12.41, SD = 2.48) compared to those wearing it on their non-dominant wrist (*M* = 13.58, SD = 1.03; *t*(58) = 2.25, *p* = 0.028). No differences according to wrist were observed for any remaining measures, including for reporting of problems wearing the accelerometer. As a result, wear wrist was not included as a covariate in the analyses below.

At least half of the participants in both age groups reported experiencing problems with the accelerometer: all these pupils reported some (usually mild) discomfort and/or inconvenience wearing the device. Only one (primary) pupil reported having problems completing the questionnaire(s), but did not provide any further details

### Associations of sleep characteristics with age, sex and day of week

In primary sleep characteristic analyses, mixed-effects linear regression models examined the associations of key activity and sleep variables with age, sex, and the day of week (weekday versus weekends) in the 59 participants with available data for each of these covariates (Table 3). To compare weekday versus weekend effects, average values across school nights (Sunday – Thursday) were compared with the average across weekend nights (Friday – Saturday), clustering by school and by individual.

**Table 3.** Mixed effects model associations between age, sex and day of week on key sleep/circadian outcome variables, clustering by school and individual.

|  | Age |  | Sex* |  | Weekday vs. weekend |  |
| --- | --- | --- | --- | --- | --- | --- |
|  | B (CI) | p | B (CI) | P | B (CI) | p |
| L5 | -0.15<br>(-0.30, 0.001) | 0.052 | 0.07<br>(-0.45, 0.59) | 0.788 | -0.12<br>(-0.28, 0.04) | 0.133 |
| M10 | -6.01<br>(-8.67, -3.36) | **<0.001** | 7.68<br>(3.12, 12.25) | **0.001** | -15.62<br>(-24.44, -6.80) | **0.001** |
| SPT-window | -0.27<br>(-0.34, -0.21) | **<0.001** | -0.35<br>(-0.71, 0.01) | 0.057 | 0.59<br>(0.26, 0.91) | **<0.001** |
| Sleep duration | -0.20<br>(-0.25, -0.14) | **<0.001** | -0.29<br>(-0.60, 0.01) | 0.062 | 0.49<br>(0.29, 0.69) | **<0.001** |
| SPT-window meets recommended sleep duration* | 1.06 (0.61, 1.85) | 0.840 | 0.39 (0.11, 1.32) | 0.130 | 5.22 (1.80, 15.18) | **0.002** |
| Sleep efficiency | 0.01<br>(-1.4x10<sup>-3</sup>, 0.01) | 0.118 | -2.3x10<sup>-3</sup><br>(-0.02, 0.02) | 0.811 | -4.7x10<sup>-3</sup><br>(-0.01, 4.9x10<sup>-3</sup>) | 0.337 |
| Sleep onset (accelerometry) | 0.19<br>(0.05, 0.33) | **0.007** | -0.10<br>(-0.85, 0.65) | 0.792 | 0.87<br>(0.70, 1.04) | **<0.001** |
| Wake time (accelerometry) | -0.09<br>(-0.26, 0.08) | 0.310 | -0.49<br>(-0.92, -0.05) | **0.029** | 1.36<br>(1.05, 1.66) | **<0.001** |
| Subjective sleep onset | 0.18<br>(-0.30, 0.66) | 0.457 | -0.17<br>(-0.63, 0.29) | 0.470 | 1.19<br>(0.50, 1.89) | **0.001** |
| Subjective wake time | -0.16<br>(-0.32, 2.6x10<sup>-3</sup>) | 0.054 | -0.32<br>(-0.75, 0.12) | 0.159 | 2.14<br>(1.91, 2.36) | **<0.001** |
| Subjective sleep duration | -0.31<br>(-0.85, 0.23) | 0.262 | -0.17<br>(-0.27, -0.07) | **0.001** | 0.94<br>(0.47, 1.42) | **<0.001** |

N = 59 as sex was missing for two participants. Day: reference category = weekdays. Sex: reference category = F. \*Coefficients are linear mixed model coefficients and 95% CI for all variables apart from 'SPT-window meets recommended sleep duration' where coefficients are odds ratios (95% CI) from mixed-effects logistic regression CI = confidence interval.

Older age was associated with lower activity levels during the most active 10 hours (M10, typically daytime activity), with shorter time in bed (SPT-window), shorter objective sleep duration and later sleep onset (Table 3).

Boys showed higher daytime (M10) activity than girls, were more likely to wake earlier (as measured by accelerometry but not by subjective reports), and to subjectively report shorter sleep duration.

Daytime (M10) activity was lower on weekends versus weekdays. At weekends, participants also spent longer in bed, had longer sleep duration, and went to bed and woke later, according to both objective and subjective measures of sleep. The odds of the sleep window duration meeting age-specific recommendations for sleep duration (Paruthi et al., 2016) were higher for weekends vs. weekdays.

### Summary and comparison of sleep characteristics for primary and secondary pupils

Primary and secondary pupils were compared using Independent t-tests (continuous measures) and chi-square tests (categorical measures) (Table 2). Primary and secondary pupils had an average sleep duration of less than 8 hours per night (7.7 hours and 7.3 hours, respectively), with only one (secondary) pupil meeting age-specific sleep duration recommendations (Paruthi et al., 2016). SPT-window duration (time in bed) was shorter for secondary (8.4 hours) than for primary (8.8 hours) pupils, and only around a third of both age groups met sleep duration recommendations.

Secondary compared to primary pupils demonstrated later onset of sleep on weekdays, lower overall levels of activity during their most active 10 hours, and lower RA (0.92 vs. 0.90).

There were no significant differences between primary and secondary pupils for the remaining objective and subjective sleep/circadian features, depressive symptoms (MFQ), or ASWS sleep quality measures.

For illustrative purposes, rest-activity patterns for a pupil showing relatively good sleep duration, sleep efficiency and RA, and a pupil showing lower values on these metrics (indicative of poorer sleep quality and duration) are depicted in Figure 1.

**Figure 1.**
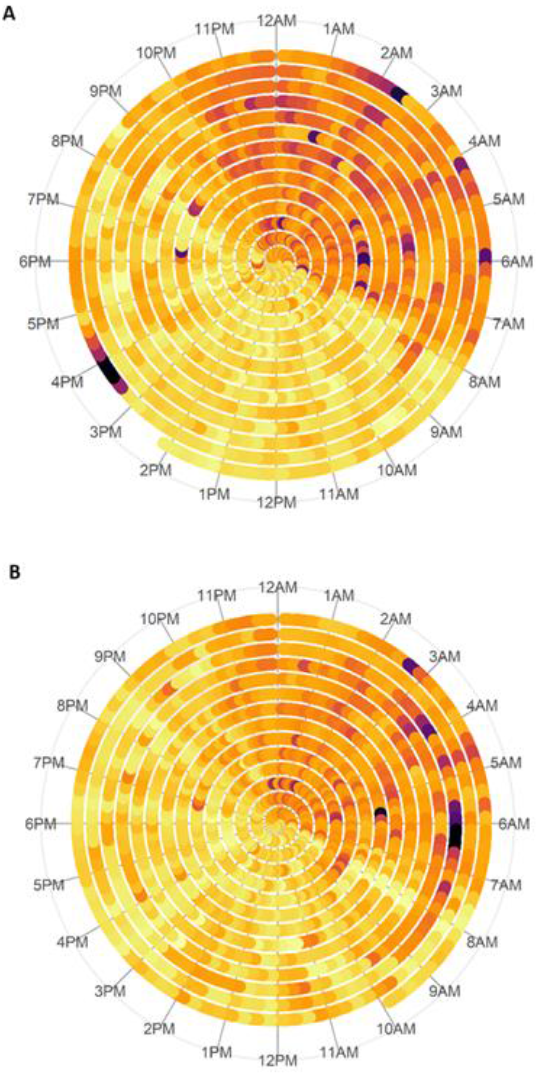
Visual representation of rest-activity patterns over 14-day data collection period. Darker colours (orange, purple) represent low activity levels; lighter colours (yellow, light orange) represent higher activity levels. A) Shows activity data from a participant with good sleep duration, RA and sleep efficiency; B) shows activity for a participant with shorter sleep duration and lower RA and sleep efficiency.

### Associations of sleep characteristics with depressive symptoms

Unadjusted pairwise correlations were examined between key objective and subjective sleep and circadian measures with depressive symptoms (Table 4).

**Table 4.**
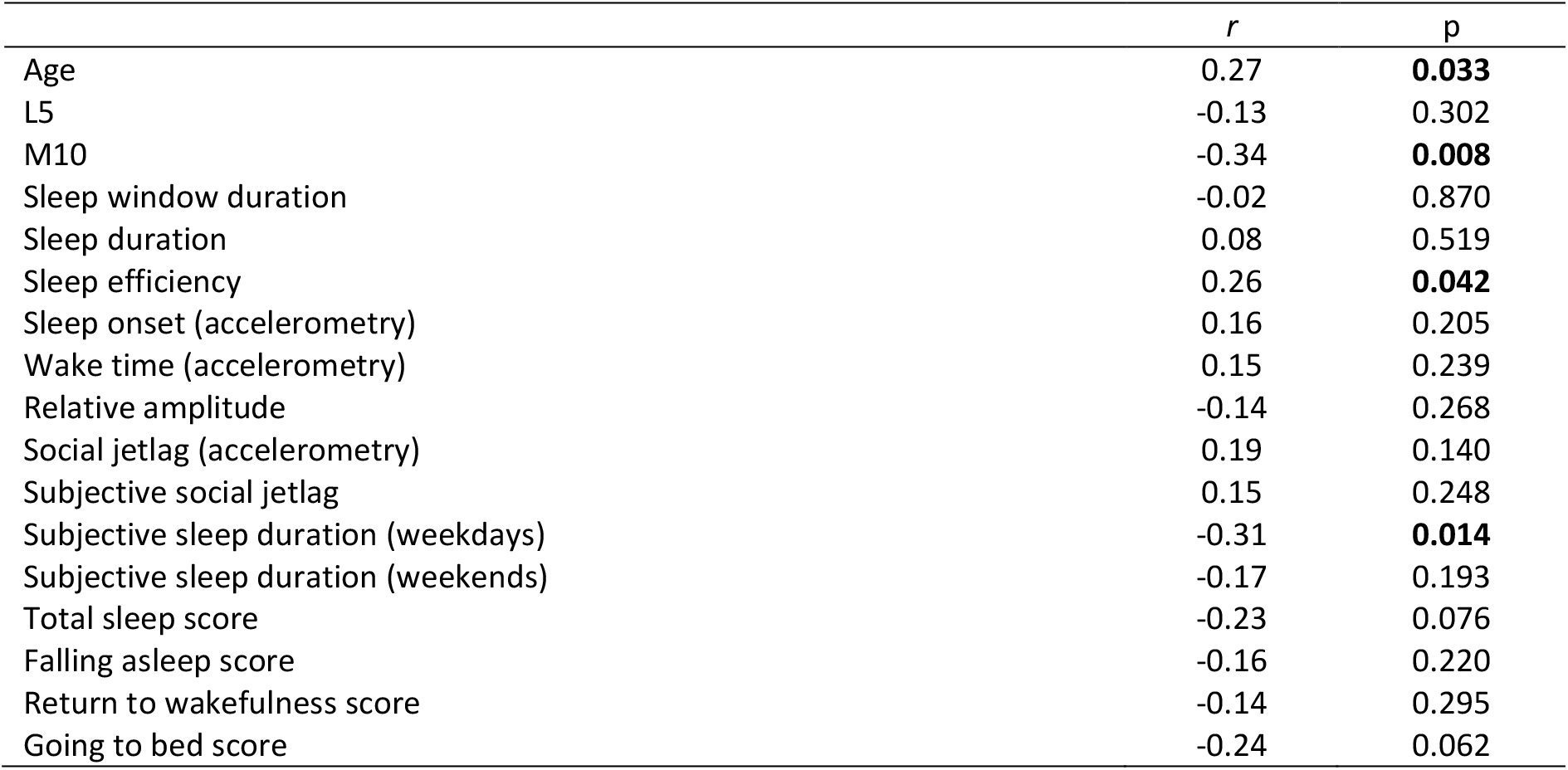
Pairwise correlations of sleep/circadian characteristics with depressive symptom scores (MFQ).

|  | *r* | *p* |
| --- | --- | --- |
| Age | 0.27 | **0.033** |
| L5 | -0.13 | 0.302 |
| M10 | -0.34 | **0.008** |
| Sleep window duration | -0.02 | 0.870 |
| Sleep duration | 0.08 | 0.519 |
| Sleep efficiency | 0.26 | **0.042** |
| Sleep onset (accelerometry) | 0.16 | 0.205 |
| Wake time (accelerometry) | 0.15 | 0.239 |
| Relative amplitude | -0.14 | 0.268 |
| Social jetlag (accelerometry) | 0.19 | 0.140 |
| Subjective social jetlag | 0.15 | 0.248 |
| Subjective sleep duration (weekdays) | -0.31 | **0.014** |
| Subjective sleep duration (weekends) | -0.17 | 0.193 |
| Total sleep score | -0.23 | 0.076 |
| Falling asleep score | -0.16 | 0.220 |
| Return to wakefulness score | -0.14 | 0.295 |
| Going to bed score | -0.24 | 0.062 |

Higher daytime activity (M10) and higher subjectively-reported sleep duration on weekdays were associated with lower depressive symptom scores. Greater (accelerometry-derived) sleep efficiency was correlated with *higher* depressive symptom scores, as was older age.

### Comparison of objective and subjective sleep/circadian measures

Accelerometry-derived and subjective reports of sleep onset and wake times, sleep window duration (both weekdays and weekends) and social jetlag estimates were compared via paired *t-*tests and pairwise correlations (Table 5). As subjective sleep/wake times were reported categorically from a finite range of 30-minute windows rather than as continuous times, direct comparisons of objective and subjective sleep timing measures below should be interpreted with caution.

**Table 5.** Comparison of subjective and objective sleep/circadian measures.

| *M* (SD) | Accelerometry | Subjective | *t* | *p* | *R* | *p* |
| --- | --- | --- | --- | --- | --- | --- |
| Sleep onset – weekdays | 22:37 (50.9) | 22:07 (50.4) | 3.95 | **<0.001** | 0.33 | **0.009** |
| Wake time – weekdays | 07:02 (38.8) | 07:07 (34.4) | 1.06 | 0.292 | 0.62 | **<0.001** |
| Sleep onset – weekends | 23:30 (71.3) | 23:24 (89.0) | 0.46 | 0.647 | 0.16 | 0.220 |
| Wake time – weekends | 08:24 (72.9) | 09:16 (86.2) | 4.06 | **<0.001** | 0.22 | 0.082 |
| Sleep window duration – weekdays | 8.45 (1.14) | 8.93 (1.14) | 2.89 | **0.005** | 0.42 | **<0.001** |
| Sleep window duration - weekends | 8.91 (1.23) | 9.83 (1.76) | 3.75 | **<0.001** | 0.13 | 0.310 |
| Social jetlag | 1.12 (0.84) | 1.71 (1.05) | 3.45 | **0.001** | 0.02 | 0.903 |

SD = standard deviation. Values for sleep onset and wake times are time (24hr clock), and in brackets, SD in minutes

Objective and subjective measures of SPT-window onset, wake times and total SPT-window duration were positively correlated on weekdays, but not weekends. Subjectively-reported weekday sleep onset was on average around 30 minutes earlier than objectively-derived estimates, and participants reported later weekend waking times (by 52 minutes) than those derived via accelerometry. As a result, SPT-window durations derived from subjective estimates were longer than objectively-measured sleep windows for weekends and weekdays.

### Associations with school deprivation level

Supplementary Table 1 reports comparison of sleep and questionnaire measures between pupils from schools in areas of low and high deprivation, based on SIMD. An important caveat is that the accelerometry data collection period overlapped with the end of DST for the higher deprivation schools (n = 24), but not the low deprivation schools. Accelerometry data were adjusted for this time change (see *Accelerometry pre-processing*), but it remains possible that differences in between schools of high and low deprivation were linked to the DST change rather than/in combination with the SIMD difference.

A greater proportion of pupils from deprived schools were excluded from analyses due to providing insufficient valid accelerometry data (high deprivation: 0.23, n = 7; low deprivation: 0.03, n = 1; Supplementary Table 1), but of those meeting data quality inclusion criteria, the number of days/nights of valid data did not differ. Pupils from more affluent compared to more deprived schools had longer subjective sleep duration and earlier subjective sleep onset (Supplementary Table 1), as well as higher self-rated Total Sleep Scores (ASWS). Objective sleep measures did not differ between these groups. More affluent school pupils had significantly lower relative amplitude.

## Discussion

Our results are consistent with previous findings that the majority of schoolchildren obtain insufficient sleep and suffer from desynchrony between their internal circadian clock and the *social clock* (Wheaton et al., 2018; Wittmann et al., 2006). We have demonstrated the feasibility of recruiting schools and pupils to accelerometry-based research into the role of sleep in mental wellbeing: the pupil response rate and data completeness were good, with 82% of participants providing sufficient accelerometer and questionnaire data for inclusion. Future investigations capitalising on the SHINE network to recruit larger and more representative samples of schoolchildren will work to establish the factors associated with such sleep and circadian disturbances, and their impact on mental health

### Feasibility

Compliance with the study instructions was good: 61/69 pupils who were provided with an accelerometer provided sufficient data for inclusion. However, the study highlighted areas for improvement in future investigations. Of the target sample of 100 pupils, 61 were recruited and provided sufficient accelerometry and questionnaire data for inclusion. Response rates for consent were very good for two schools (80-100%), but were lower than expected at the two other schools where less than 60% of the targeted 25 pupils agreed to participate (Table 1). Response rates in future may benefit from greater public engagement activities to highlight the importance of sleep and mental health research prior to recruitment –at one school where such an activity took place before study recruitment, 100% of the targeted 25 pupils were recruited, and of these all but one (96%) provided sufficient data for inclusion.

Over half of participants reported some discomfort/inconvenience wearing the accelerometer. Feedback suggested this was due to the size and fit of the wristband: this discomfort likely contributed to the exclusion of 8 pupils who did not have sufficient wear-time for inclusion. Future studies could avoid this issue by using wristbands specifically designed for children.

Pupils were asked to wear the accelerometer on their dominant wrist, but only 56% did so, and those who did provided on average fewer nights of valid data than those wearing the device on their non-dominant wrist. This may suggest greater discomfort/inconvenience due to the device on the dominant wrist, although this was not borne out by increased reporting of problems with the accelerometer in the feedback questionnaire. Consistent with previous studies, no other group differences according to wrist were observed (Dieu et al., 2017; Driller, O’Donnell, & Tavares, 2017). Future investigations may achieve greater compliance and data completeness by advising participants to use their non-dominant wrist.

### Sleep duration and efficiency

A 2016 American Academy of Sleep Medicine (AASM) consensus statement recommended that for optimal health children aged 6-12 years should regularly sleep 9-12 hours per night, and teenagers 8-10 hours (Paruthi et al., 2016). In our study primary schoolchildren (aged around 11 years) slept for 7.7 hours on average, and none had an average sleep duration within the recommended 9-12 hour range (Table 2). Secondary schoolchildren had an average sleep duration of 7.4 hours, and only one had an average sleep duration within the recommended range. Even when considering accelerometry-derived sleep window time, two thirds of children did not allow enough time in bed to obtain sufficient sleep. These findings are concerning, but not unusual in the context of previous large-scale studies (Chaput & Janssen, 2016; Wheaton et al., 2018).

Older pupils were particularly prone to short sleep duration, spending less time in bed, sleeping for a shorter time, and going to bed later on weekdays (Tables 2 and 3). Older age was also associated with lower levels of daytime (M10) activity. Possible explanations may include older children experiencing less parental control of sleep times (Meijer, Habekothé, & Van Den Wittenboer, 2001), greater pre-bedtime technology use (Harbard et al., 2016), or greater school workload (Galloway, Conner, & Pope, 2013). These factors, alongside the age-related circadian shift, may result in greater interference with sleep timing compared to younger children (Wittmann et al., 2006). Older pupils did not however show greater social jetlag effects or differences in weekend wake times. Social or parental constraints may necessitate earlier rising on weekends than young adolescents would naturally prefer, compounding effects of shifting chronotype.

The AASM recommendations do not take account of interindividual variation in the need for sleep, and other measures of sleep quality are also likely to be important (Gruber et al., 2014). Sleep efficiency of > 85% has been considered to represent good sleep quality (Ohayon et al., 2017), and primary and secondary pupils in our study exceeded this, spending around 87% of time in bed asleep. RA values were also indicative of relatively healthy rest-activity rhythms, with average values (0.90 and 0.92 for primary and secondary pupils, respectively) exceeding those for a large population-based sample of middle-aged UK adults (Lyall et al., 2018).

These data therefore give a mixed picture as to overall sleep quality (duration and efficiency), but differences in many sleep characteristics between weekdays and weekends are evident. Participants showed a difference in sleep timing (midpoint of sleep) of over an hour between school nights and weekends: this *social jetlag* effect is not excessive (Wittmann et al., 2006), but is indicative of a degree of desynchrony between the biological circadian clock and the social clock enforced by school timings. Consistent with reverting to the body’s natural rhythms and catching up on sleep lost during the week, on weekends most participants went to bed later, spent longer in bed, slept for longer and woke later.

### Associations with depressive symptoms

Previous evidence suggests a link between poor sleep quality, shorter sleep duration and worse mental health and wellbeing in children and adolescents, including more depressive symptoms (Lovato & Gradisar, 2014). Consistent with this, longer subjective sleep duration was associated with lower depressive symptoms. However, greater objectively-measured sleep efficiency was associated with *greater* depressive symptoms. Scores on the MFQ were on average very low (3.8 for primary and 5.4 for secondary pupils, out of 26), and well below the cut-off of 12 indicating possible depression. This, combined with the sample’s high overall sleep efficiency may have resulted in a spurious association due to floor and/or ceiling effects for these variables, respectively.

Higher daytime activity levels were correlated with lower MFQ scores. This is consistent with meta-analytic and prospective cohort evidence (Carter, Morres, Meade, & Callaghan, 2016; Kandola, Lewis, Osborn, Stubbs, & Hayes, 2020) of associations between greater physical activity and lower depressive symptoms in adolescents. Although the primary focus of this study was sleep and circadian characteristics, the role of physical activity will be addressed further in later studies within SHINE. Further measures of subjective wellbeing and mental health will also be of use in identifying associations with sleep.

### Objective vs. subjective measures

We found moderate agreement between objective and subjective measures of sleep timing and duration. Several measures were positively correlated, but participants reported earlier weekday bedtimes and longer weekday sleep duration than was found by accelerometry. Participants also reported later waking times and longer sleep duration (by about one hour) on weekends, compared to objective measures.

Adolescents, particularly those with depression or anxiety, have been found to overestimate sleep problems relative to objective measures (Alfano, Patriquin, & De Los Reyes, 2015). Conversely however, some studies have reported a tendency for healthy children and adolescents to overestimate total sleep time, via underappreciation of waking during the night (Tremaine, Dorrian, & Blunden, 2010). This latter point is in line with our findings, where objective measures of sleep duration were typically shorter than subjective estimates.

The findings suggest that self-reported measures of sleep timing and sleep duration provide an incomplete picture of sleep characteristics and should be used alongside objective measures (Arora, Broglia, Pushpakumar, Lodhi, & Taheri, 2013).

## Conclusion

This study demonstrates the feasibility of conducting schools-based research into the links between sleep and mental health in young people in Scotland. Our data highlight potential issues of inadequate sleep and circadian desynchrony within this group, particularly in secondary school pupils. Notably, reduced daytime activity was linked to greater reporting of depressive symptoms but given the cross-sectional nature of the data, conclusions cannot be drawn as to the causal direction of this association. Future larger-scale investigations incorporating data from schools across different latitudes in Scotland - and a greater range of mental health, wellbeing and cognitive assessment tools - will facilitate a more detailed examination of the role of sleep and circadian disturbance in the mental wellbeing of schoolchildren.

## Data Availability

Data available from authors on request.

## Acknowledgements

We are grateful to the participating SHINE schools, teachers and pupils who agreed to take part in the study. This work was supported by an MRC Mental Health Data Pathfinder Award (MC_PC_17217). DJS acknowledges the support of the Brain and Behaviour Research Foundation (Independent Investigator Award 1930) and a Lister Prize Fellowship (173096). SS and JI were supported by an MRC Strategic Award (MCC_UU_12017_14 & SPHSU14). LML is supported by a JMAS Sim Fellowship for depression research from the Royal College of Physicians of Edinburgh.

---

**Supplementary Table 1.** Comparison of sample characteristics between pupils from low and high deprivation schools (SIMD)

| Measure; M (SD) | High<br>(n = 24) | Low<br>(n = 37) | t/$\chi^2$ | p |
| --- | --- | --- | --- | --- |
| Datasets excluded, N (% of sample with accelerometry) | 7 (22.58) | 1 (2.63) | 6.63 | **0.010** |
| Age* | 11.88 (1.39) | 11.57 (0.55) | 1.21 | 0.232 |
| Secondary school, N (%) | 7 (29.17) | 24 (64.86) | 7.42 | **0.006** |
| Female*, N (%) | 13 (54.17) | 26 (70.27) | 1.64 | 0.441 |
| missing, N (%) | 1 (4.17) | 1 (2.70) |  |  |
| Number valid weekdays | 8.08 (1.69) | 8.54 (1.17) | 1.25 | 0.217 |
| Number of valid weekend days | 3.58 (0.83) | 3.63 (0.86) | 0.17 | 0.864 |
| Number of nights | 12.50 (2.43) | 13.19 (1.71) | 1.30 | 0.199 |
| Wore actigraph on dominant wrist, N (%) | 16 (66.67) | 18 (48.65) | 4.07 | 0.131 |
| missing, N (%) | 1 (4.17) | 0 (0) |  |  |
| **Accelerometry-derived activity measures** |  |  |  |  |
| Activity (ENMO) during least active 5 hours (L5) | 2.82 (0.78) | 3.07 (0.77) | 1.22 | 0.229 |
| Activity (ENMO) during most active 10 hours (M10) | 70.72 (22.02) | 61.70 (18.73) | 1.71 | 0.092 |
| **Accelerometry-derived sleep measures** |  |  |  |  |
| SPT-window duration | 8.55 (0.76) | 8.64 (0.70) | 0.48 | 0.636 |
| Sleep duration | 7.48 (0.79) | 7.49 (0.66) | 0.04 | 0.966 |
| Sleep efficiency | 0.88 (0.04) | 0.87 (0.03) | 1.32 | 0.193 |
| Sleep onset – weekdays | 22:36 (57.0) | 22:38 (47.4) | 0.13 | 0.901 |
| Sleep onset – weekends | 23:31 (78.6) | 23:29 (67.2) | 0.06 | 0.954 |
| Wake time – weekdays | 07:01 (52.2) | 07:04 (27.6) | 0.35 | 0.727 |
| Wake time - weekends | 08:07 (82.2) | 08:36 (64.8) | 1.55 | 0.127 |
| Meets sleep duration recommendations, N (%) | 0 (0) | 1 (2.70) | 0.66 | 0.417 |
| SPT-window meets recommended sleep duration, N (%) | 10 (41.67) | 11 (29.73) | 0.92 | 0.338 |
| **Accelerometry-derived circadian measures** |  |  |  |  |
| Relative amplitude | 0.92 (0.02) | 0.90 (0.03) | 2.44 | **0.018** |
| Social jetlag (hours) | 1.01 (0.92) | 1.20 (0.79) | 0.86 | 0.394 |
| **Questionnaires** |  |  |  |  |
| Total sleep score | 33.54 (9.66) | 38.38 (8.40) | 2.07 | **0.043** |
| Falling Asleep & Reinitiating Sleep | 18.38 (6.86) | 21.54 (5.72) | 1.95 | 0.056 |
| Returning to Wakefulness | 5.42 (2.48) | 5.49 (2.38) | 0.11 | 0.913 |
| Going to Bed | 9.75 (3.43) | 11.35 (2.85) | 1.98 | 0.053 |
| Mood and Feelings score | 5.67 (5.12) | 3.89 (3.39) | 1.63 | 0.108 |
| **Subjective sleep/circadian measures** |  |  |  |  |
| Subjective sleep onset – weekdays | 22:35 (58.2) | 21:49 (34.8) | 3.82 | **<0.001** |
| Subjective wake time - weekdays | 06:56 (41.8) | 07:14 (27.0) | 1.99 | 0.051 |
| Subjective sleep onset – weekends | 00:24 (91.8) | 22:45 (62.4) | 5.04 | **<0.001** |
| Subjective wake time - weekends | 09:19 (100.2) | 09:14 (76.8) | 0.18 | 0.856 |
| Subjective sleep duration – weekdays | 8.35 (1.39) | 9.41 (0.75) | 3.83 | **<0.001** |
| Subjective sleep duration - weekends | 8.92 (1.87) | 10.50 (1.37) | 3.80 | **<0.001** |
| Subjective social jetlag | 2.09 (1.24) | 1.47 (0.83) | 2.37 | **0.021** |
| **Feedback questions** |  |  |  |  |
| Problems with questionnaires? N (%) | 1 (4.17) | 0 (0) | 2.84 | 0.242 |
| Problems with actigraph? N (%) | 14 (58.33) | 18 (48.65) | 1.06 | 0.588 |

\* Age and sex are estimated, see *Participants and Procedure*. For sleep onset and wake times, values are time in 24hr clock (SD in minutes). ENMO = Euclidean Norm Minus One; SIMD = Scottish Index of Multiple Deprivation; SPT = Sleep Period Time.

